# Large-language-model-based 10-year risk prediction of cardiovascular disease: insight from the UK biobank data

**DOI:** 10.1101/2023.05.22.23289842

**Authors:** Changho Han, Dong Won Kim, Songsoo Kim, Seng Chan You, SungA Bae, Dukyong Yoon

## Abstract

**Background:** Conventional cardiovascular risk prediction models provide insights into population-level risk factors and have been widely adopted in clinical practice. However, these models have limited generalizability and flexibility. Large language models (LLMs) have demonstrated remarkable proficiency for use in various industries.

**Methods:** In this study, we have investigated the feasibility of Large Language Models (LLMs) such as ChatGPT-3.5, ChatGPT-4, and Bard for predicting 10-year cardiovascular risk of a patient. We used data from the UK Biobank Cohort, a major biomedical database in the UK, and the Korean Genome and Epidemiology Study (KoGES), a large-scale prospective study in Korea, for additional validation and multi-institutional research. These databases provided a wide array of information including age, sex, medical history, lipid profile, blood pressure, and physical measurement. Based on these data, we generated language sentences for individual analysis and input these into the LLM to derive results. The performance of the LLMs was then compared with the Framingham Risk Score (FRS), a conventional risk prediction model, using this real-world data. We confirmed the model performance of both the LLMs and FRS, evaluating their accuracy, sensitivity, specificity, Positive Predictive Value (PPV), Negative Predictive Value (NPV), and F1 score. Their performance in predicting 10-year cardiovascular risk was compared through Kaplan-Meier survival analysis and Cox-hazard ratio analysis.

**Findings:** GPT-4 achieved performance comparable to the FRS in cardiovascular risk prediction in both the UK Biobank {accuracy (0·834 vs· 0·773) and F1 score (0·138 vs· 0·132)} and KoGES {accuracy (0·902 vs· 0·874)}. The Kaplan–Meier survival analysis of GPT-4 demonstrated distinct survival patterns among groups, which revealed a strong association between the GPT risk prediction output and survival outcomes. The additional analysis of limited variables using GPT-3·5 indicated that ChatGPT’s prediction performance was preserved despite the omission of a few variables in the prompt, especially without physical measurement data

**Interpretation:** This study proposed that ChatGPT can achieve performance comparable to conventional models in predicting cardiovascular risk. Furthermore, ChatGPT exhibits enhanced accessibility, flexibility, and the ability to provide user-friendly outputs. With the evolution of LLMs, such as ChatGPT, studies should focus on applying LLMs to various medical scenarios and subsequently optimizing their performance.

## Introduction

Cardiovascular disease (CVD) is the leading cause of morbidity and mortality worldwide, accounting for a considerable proportion of healthcare costs and posing a substantial public health risk^1^. The accurate and timely prediction of an individual’s risk of developing CVD can facilitate early intervention and prevention strategies, which reduces the incidence and impact of CVD^2^. Although conventional CVD risk prediction models, such as the Framingham risk score (FRS)^3^, American College of Cardiology/American Heart Association (ACC/AHA) Pooled Cohort Equations^4^, and the QRISK3 score^5^ provide insights into population-level risk factors and have been widely adopted in clinical practice, these models have several limitations. First, these models have limited generalizability to diverse populations with varying demographic, clinical, and genetic characteristics^6^. Second, conventional models may not incorporate novel risk factors or consider complex interactions between risk factors, leading to potential underestimation or risk overestimation^7^. Third, the implementation of conventional risk models can be challenging because of complex calculations and variable requirements, which hinders their use in clinical settings^8^. Finally, these risk models do not have adequate personalization and rely on population-level data, which may not accurately capture individual-level variations in risk factors. This phenomenon limits their ability to provide tailored risk assessments for patients^9^.

Advances in artificial intelligence (AI) can overcome these limitations^10^. Large language models (LLMs), particularly the generative pretrained transformer 4 (GPT-4) model developed by OpenAI, exhibit remarkable proficiency in producing human-like languages, and have potential for application in other industries^11, 12^. However, despite their growing popularity, reliability concerns severely affect the use of LLMs in the medical field, which requires precise and accurate information. A potential problem with LLMs is “AI hallucination.” This phenomenon occurs when AI confidently generates an impressive-sounding response that may not be justified by its training data or may even be factually incorrect^13^. The presence of AI hallucinations raises reliability and accuracy concerns on information produced by these models, particularly in domains such as medicine, which requires precise and trustworthy information. Although such problems have been reduced in GPT4^12, 14^, only a few studies have quantified or analyzed this topic.

Although the use of language models in the medical field has attracted considerable attention, limited quantitative evaluation of their performance and accuracy has been conducted in specific medical tasks^15^. Therefore, we evaluated LLMs in predicting CVD risk for 10 years and compared their performance with that of conventional risk prediction models using UK Biobank and Korean Genome and Epidemiology Study (KoGES) data^16, 17^.

## Methods

### Data source and outcome assessment

We used data from the UK Biobank cohort, a large-scale biomedical database of UK general population. Established in 2006, the UK Biobank cohort is one of major international health resources that has collected extensive data and biological samples from approximately 500,000 participants aged between 40 and 69 years at the time of assessment. We used the UK Biobank database to extract data pertaining to age, sex, diabetes diagnosed by a doctor, blood pressure medication, smoking status, total cholesterol, high density lipoprotein (HDL) cholesterol, low density lipoprotein (LDL) direct, triglycerides, systolic blood pressure, diastolic blood pressure, standing height, weight, date of attending the assessment center, and date of death (Supplementary Table 1). We excluded cases with any missing values among these variables, except death date and ethnicity. After exclusion, 10,000 individuals were selected through random sampling. Additionally, individuals who previously experienced major cardiovascular adverse events (MACE) were excluded from the study (Fig. 1).

**Fig. 1.**
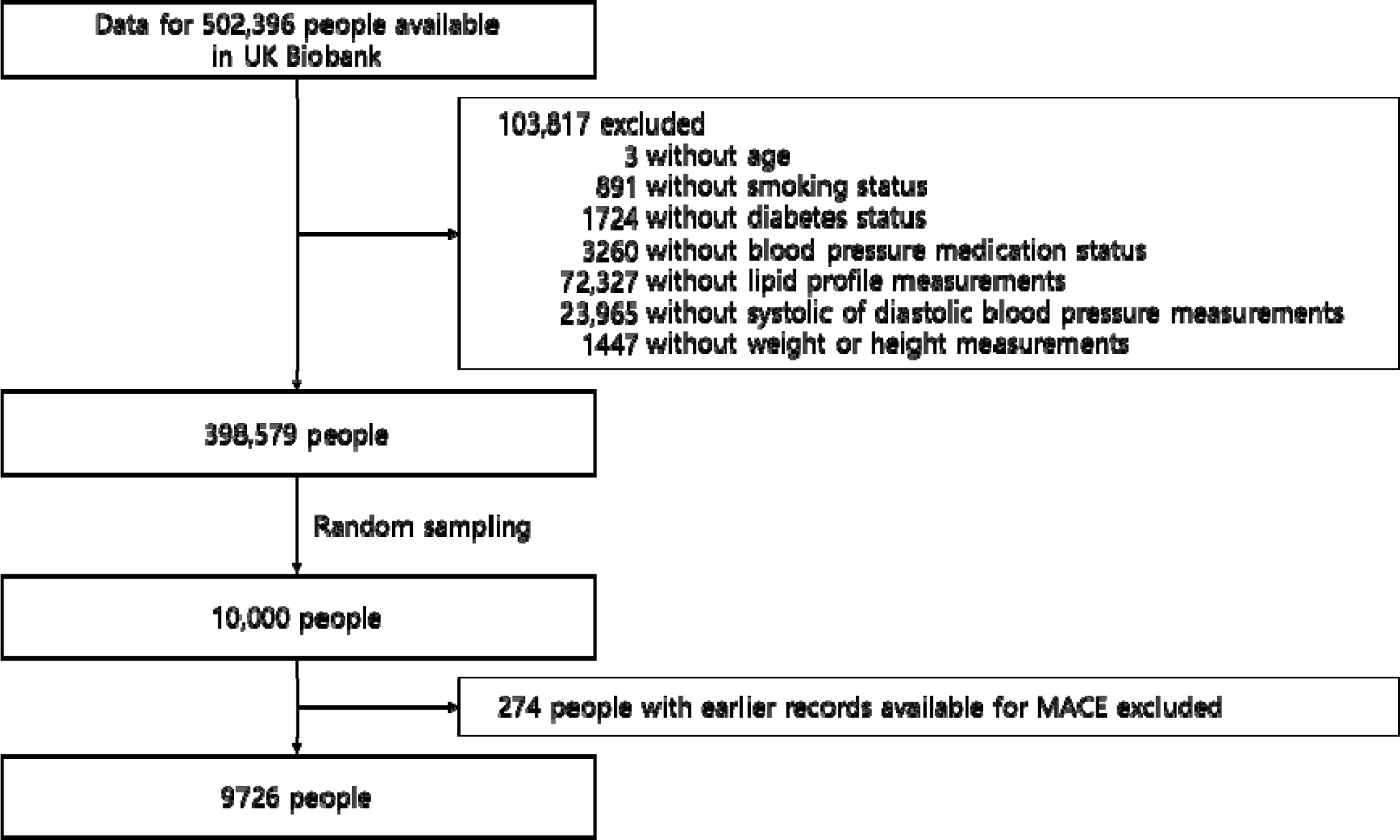
Selection of study population. MACE: major adverse cardiovascular events.

The FRS was originally developed for predicting coronary heart disease (CHD) but has since been evolved for use in forecasting not only CHD but also cerebrovascular disease, peripheral artery disease (PAD), and heart failure (HF)^3^. Using this approach, we assessed patient outcomes using MACE, which represents the most fatal and predominant occurrence of CVD. MACE is defined as follows: it was defined by the earliest recorded event of myocardial infarction (International Classification of Diseases [ICD]9 codes 410, 411·0, 412, 429·79, or ICD10 codes I21, I22, I23, I24.1, I25.2 or UK Biobank Self Report field 20002 codes 1075) or ischemic stroke (ICD9 codes 434, 436 or ICD10 codes I63, I64 or UK Biobank Self Report field 20002 codes 1583)^18^. The outcome was obtained through a category called an algorithmically defined outcome, which included information on the likely instances of specific health issues, derived from the algorithmic integration of coded data from the UK Biobank’s initial assessment data compilation (incorporating data from participants regarding their self-reported medical conditions, surgeries, and medications), in conjunction with associated data from hospital admissions (diagnoses and procedures) and death records.

In addition to the UK Biobank dataset, we used KoGES data as an additional validation cohort. The KoGES is a large-scale prospective study designed to investigate the genetic and environmental factors contributing to chronic diseases in the Korean population^16^. We used baseline data from the KoGES cohort collected between 2001 and 2002 to extract variables analogous to those used in the UK Biobank. These variables included age, sex, diagnosis of diabetes by a physician, blood pressure medication use, smoking status, total cholesterol, HDL, triglycerides, systolic blood pressure, diastolic blood pressure, height, and weight. LDL levels were calculated based on total cholesterol, HDL, and triglyceride levels (Supplementary Table 2). In this cohort, we defined patients as those who experienced a disease event (myocardial infarction or cerebrovascular disease) at least once during the 10-year follow up, resulting in 176 patients. Detailed inclusion and exclusion criteria on KoGES population selection are described in Supplementary figure 1.

### Cardiovascular risk calculation—the conventional score (Framingham Risk Score, FRS)

The FRS is a widely recognized and well-established algorithm that is used to estimate an individual’s 10-year risk of developing CVD.^3^ This score considered various factors, including age, sex, blood pressure, cholesterol levels, smoking status, and diabetes. The details are listed in Supplementary Table 1 and Supplementary Table 2. In this study, we used the FRS to calculate the cardiovascular risk percentage for each individual. Based on these percentages, we classified their risks into distinct categories (low, moderate, or high) to facilitate a comprehensive understanding of their potential for developing CVD.

### Cardiovascular risk calculation—LLMs (GPT-3·5/GPT-4/bard)

To predict the incidence of CVDs using LLMs (ChatGPT and Bard), we reformatted the variables into a sentence structure, as detailed in Fig. 2, prior to feeding them into LLMs. The decision to use this conversion was based on the inherent language model nature of LLMs. This approach allowed us to specify our output to represent individual risk percentages rather than extensive text narratives. Furthermore, the approach enabled the systematic classification of risk into low, moderate, and high categories within the provided output. Information on each participant (age, sex, diabetes, hypertension, smoking status, total cholesterol, LDL cholesterol, HDL cholesterol, triglycerides, systolic blood pressure, diastolic blood pressure, and body mass index [BMI, calculated from height and weight]) was provided to the LLMs, and the 10-year CVD risk percentage was extracted using regular expressions from the corresponding answers. Based on the 10-year CVD risk percentage, 10% or less was classified as low risk, 10%–20% as moderate risk, and > 20% as high risk.

**Fig. 2.**
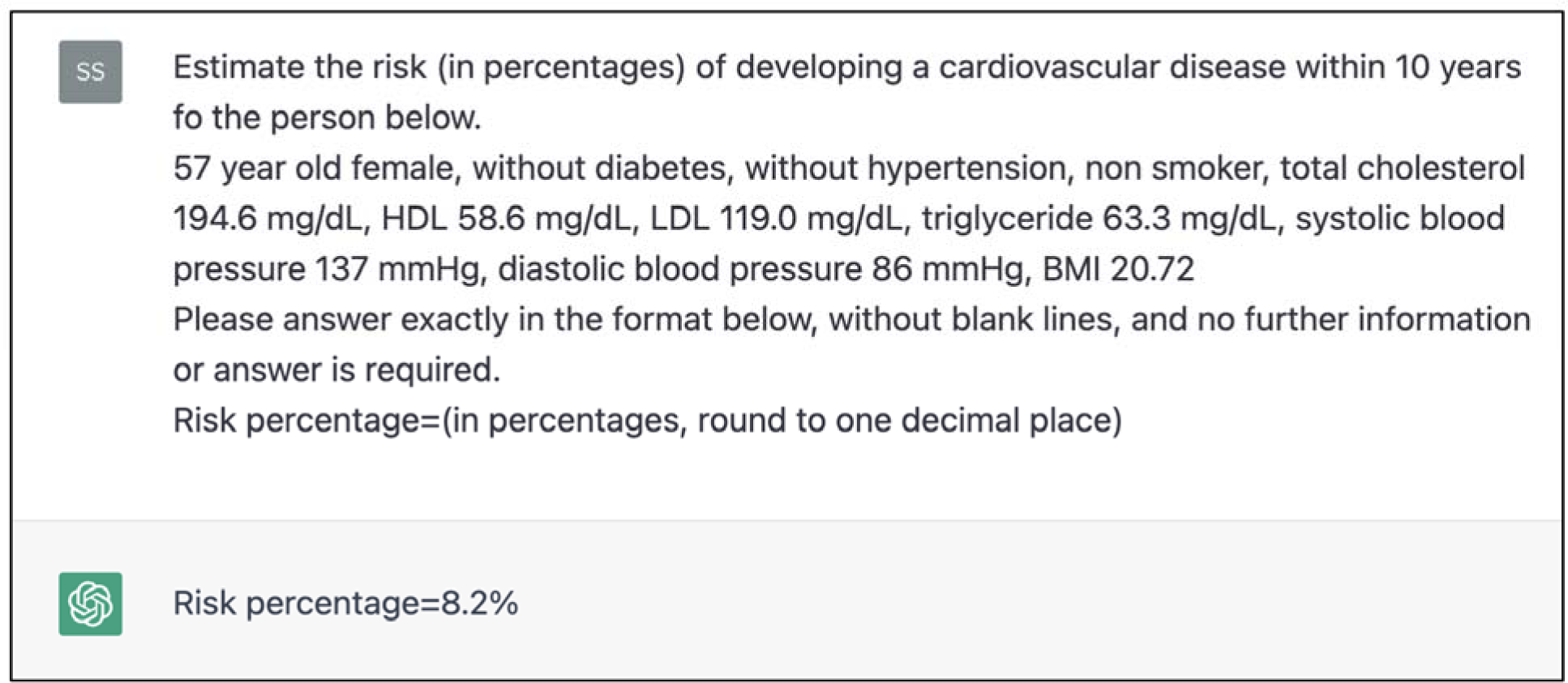
Example of a ChatGPT prompt and response for risk stratification. Tabular data extracted from the UK biobank and KoGES were organized and queried into a sentence format like the example above. The 10-year CVD risk percentage was extracted using regular expressions from the corresponding answers.

For GPT-3·5, we used the OpenAI ChatGPT API (GPT-3.5-turbo, March 23 version) in a Python environment to streamline the extraction of results. However, in the cases of GPT-4 and Bard, the lack of open-source APIs comparable to GPT-3·5 necessitated an alternative approach. GPT-4 and Bard enable accounts operating in online environments to iteratively input data and generate output text for each new chat instance.

### Model comparison between scoring systems

In the proposed methodology, the statistical significance of ChatGPT-3·5, ChatGPT-4, Bard, and FRS was investigated by calculating Pearson correlation coefficients. This numerical measure was used to assess the linearity between the output of these models and the observed data, providing an indication of both the strength and direction of these relationships.

### Performance evaluation

To assess the performance of each model, we calculated sensitivity, specificity, positive predictive value (PPV), negative predictive value (NPV), and F1 score. Sensitivity details the ability of the model to correctly identify true-positive cases, whereas specificity evaluates the accuracy of the model in identifying true-negative cases. PPV represents the proportion of true-positive cases among predicted positives, whereas NPV denotes the proportion of true-negative cases among predicted negatives. The F1 score is the harmonic mean of sensitivity and PPV, which provides a single metric for model performance, particularly in situations with imbalanced class distributions.

### LLM model performance using limited information

To evaluate the robustness of the LLMs in an environment where all input data cannot be investigated, we conducted additional experiments by constructing prompts using limited information and then querying GPT-3·5 with the UK Biobank cohort. This evaluation involved the use of an-omitting section, in which particular categories of patient data were excluded from the existing prompt. In the first experiment, data related to patient history (history of diabetes, blood pressure medication, and smoking) were excluded. In the second experiment, the data related to lipid profiles (total cholesterol, HDL cholesterol, LDL cholesterol, and triglycerides) were excluded. In the third experiment, data related to physical measurements (blood pressure and BMI) were excluded. These defined groupings are the foundation for analyzing the model performance under various conditions.

### Statistical analysis

To assess the statistical significance of differences in baseline characteristics among the risk groups, first, we performed a normality test using the Shapiro–Wilk method. After determining that our data did not satisfy the conditions of normality, we proceeded with nonparametric tests. We used the chi-squared test and Kruskal–Wallis test. By incorporating these tests into our analysis, we evaluated the statistical significance of the differences in baseline characteristics among the risk groups. Here, P-value < 0·05 was considered as significant in all tests.

The Kaplan–Meier method was applied to plot survival curves for the low-, moderate-, and high-risk groups based on 10-year mortality data from the UK Biobank. Furthermore, we used the Cox proportional hazards model to compare the survival function associated with MACE within each risk group in both the LLMs and FRS. The predictors in the model were the risk categories (low, moderate, and high) derived from the scoring systems, with the low-risk group used as a reference.

## Results

The UK Biobank study included 502,396 participants aged 40–69 years at the time of assessment, recruited between 2006 and 2010 (Fig. 1). A total of 103,817 participants with missing data were excluded. Of the remaining participants, after randomly selecting 10,000 participants, 274 patients who had previously experienced MACE (Major Adverse Cardiovascular events) were excluded, leaving 9726 subjects for the analysis.

Table 1 shows the baseline characteristics of the participants and the cardiovascular risk scores derived from the LLMs when grouped by GPT-4 category. Among a total of 9726 individuals for analysis, the participants had an overall median age of 58 years (IQR 50–63) with 4359 (44·8%) men and 331 (3·4%) experienced MACE within 10 years. When grouped by the GPT-4 category, 4222 individuals were classified as low-risk, 3957 as moderate risk, and 1547 as high risk. The higher-risk group had older individuals, a higher proportion of men, higher incidence of 10-year MACE, more diabetes mellitus, received more antihypertensive treatment, smoked more, more unfavorable lipid profiles, and higher blood pressure and BMI (all p < 0·001).

**Table 1.** Baseline characteristics (grouped by ChatGPT-4.0 category) and derived cardiovascular risk scores from large language models (LLMs)

|  | Low risk<br>(n = 4222) | Moderate risk<br>(n = 3957) | High risk<br>(n = 1547) | Overall<br>(n = 9726) | P-Value |
| --- | --- | --- | --- | --- | --- |
| Age | 52.0 [46.0-58.0] | 60.0 [54.0-64.0] | 63.0 [59.0-66.0] | 58.0 [50.0-63.0] | <0.001 |
| Sex |  |  |  |  |  |
| Female | 3015 (71.4%) | 1835 (46.4%) | 517 (33.4%) | 5367 (55.2%) | <0.001 |
| Male | 1207 (28.6%) | 2122 (53.6%) | 1030 (66.6%) | 4359 (44.8%) | <0.001 |
| Smoking status |  |  |  |  |  |
| Current | 3989 (94.5%) | 3473 (87.8%) | 1249 (80.7%) | 8711 (89.6%) | <0.001 |
| Previous, never | 233 (5.5%) | 484 (12.2%) | 298 (19.3%) | 1015 (10.4%) | <0.001 |
| Height, cm | 166.0 [161.0-173.0] | 169.0 [162.0-176.0] | 171.0 [164.0-177.0] | 168.0 [162.0-175.0] | <0.001 |
| Weight, kg | 71.0 [62.5-81.5] | 78.9 [69.6-89.2] | 84.6 [74.6-96.2] | 76.4 [66.5-87.5] | <0.001 |
| BMI, mg/kg<sup>2</sup> | 25.3 [22.9-28.2] | 27.3 [24.9-30.3] | 29.0 [26.1-32.9] | 26.7 [24.1-29.9] | <0.001 |
| Total cholesterol, mmol/L | 5.5 [4.9-6.2] | 5.9 [5.1-6.7] | 5.6 [4.6-6.6] | 5.7 [5.0-6.5] | <0.001 |
| HDL, mmol/L | 1.5 [1.3-1.8] | 1.4 [1.2-1.6] | 1.2 [1.1-1.5] | 1.4 [1.2-1.7] | <0.001 |
| LDL, mmol/L | 3.4 [2.9-3.9] | 3.7 [3.1-4.3] | 3.6 [2.8-4.3] | 3.5 [3.0-4.1] | <0.001 |
| Triglyceride, mmol/L | 1.2 [0.9-1.7] | 1.6 [1.2-2.3] | 1.9 [1.4-2.7] | 1.5 [1.0-2.1] | <0.001 |
| SBP, mm Hg | 128.5 [118.5-138.5] | 140.5 [130.0-152.5] | 150.0 [137.5-163.5] | 136.0 [124.5-149.0] | <0.001 |
| DBP, mm Hg | 79.0 [73.0-85.0] | 84.0 [77.5-90.5] | 86.5 [79.5-93.5] | 82.0 [75.5-89.0] | <0.001 |
| Antihypertensive treatment | 239 (5.7%) | 904 (22.8%) | 743 (48.0%) | 1886 (19.4%) | <0.001 |
| Diabetes mellitus | 7 (0.2%) | 89 (2.2%) | 376 (24.3%) | 472 (4.9%) | <0.001 |
| 10-year MACE | 54 (1.3%) | 147 (3.7%) | 130 (8.4%) | 331 (3.4%) | <0.001 |
| ChatGPT-4 score | 6.2 [4.1-8.3] | 14.3 [11.9-16.4] | 23.7 [21.4-27.4] | 11.3 [6.8-16.8] | <0.001 |
| Framingham |  |  |  |  |  |
| Low risk | 3327 (78.8%) | 887 (22.4%) | 41 (2.7%) | 4255 (43.7%) | <0.001 |
| Moderate risk | 814 (19.3%) | 2039 (51.5%) | 401 (25.9%) | 3254 (33.5%) |  |
| High risk | 81 (1.9%) | 1031 (26.1%) | 1105 (71.4%) | 2217 (22.8%) |  |
| Score | 6.2 [3.9-9.4] | 14.8 [10.5-20.4] | 26.7 [18.9-35.1] | 11.4 [6.4-19.1] | <0.001 |
| ACC/AHA |  |  |  |  |  |
| Low risk | 3827 (90.6%) | 1818 (45.9%) | 191 (12.3%) | 5836 (60.0%) | <0.001 |
| Moderate risk | 389 (9.2%) | 1952 (49.3%) | 835 (54.0%) | 3176 (32.7%) |  |
| High risk | 6 (0.1%) | 187 (4.7%) | 521 (33.7%) | 714 (7.3%) |  |
| **Score** | 2·4 [1·2-4·4] | 8·1 [5·0-12·5] | 16·3 [10·5-22·4] | 5·7 [2·5-11·3] | <0·001 |
| **ChatGPT-3·5** |  |  |  |  |  |
| **Low risk** | 2721 (64·4%) | 764 (19·3%) | 50 (3·2%) | 3535 (36·3%) | <0·001 |
| **Moderate risk** | 1091 (25·8%) | 1562 (39·5%) | 303 (19·6%) | 2956 (30·4%) |  |
| **High risk** | 410 (9·7%) | 1631 (41·2%) | 1194 (77·2%) | 3235 (33·3%) |  |
| **Score** | 7·6 [5·1-13·1] | 17·4 [11·8-25·1] | 27·9 [20·9-38·2] | 14·3 [7·4-23·5] | <0·001 |
| **Bard** |  |  |  |  |  |
| **Low risk** | 2131 (50·5%) | 767 (19·4%) | 128 (8·3%) | 3026 (31·1%) | <0·001 |
| **Moderate risk** | 1478 (35·0%) | 1755 (44·4%) | 605 (39·1%) | 3838 (39·5%) |  |
| **High risk** | 613 (14·5%) | 1435 (36·3%) | 814 (52·6%) | 2862 (29·4%) |  |
| **Score** | 10·0 [3·4-15·3] | 16·4 [10·5-24·3] | 20·6 [14·4-29·2] | 13·7 [7·4-22·0] | <0·001 |

Data are median (IQR) or n (%).
BMI: body mass index, SBP: systolic blood pressure, DBP: diastolic blood pressure, MACE: major adverse cardiovascular events.

Table 2 presents the performance comparison of the scores (derived from the GPT-4, GPT-3·5, Bard, and Framingham risk scores) in predicting 10-year MACE. The 10-year MACE performance prediction for people classified as high risk in each scoring system were detailed. In the UK Biobank cohort, theGPT-4 score had the highest accuracy of 0·834, specificity of 0·849, PPV of 0·084, and F1 score of 0·849, 0·084, and 0·138, respectively. The GPT-3·5 score had the highest sensitivity (0·598) and NPV (0·980). Overall, the performance of the GPT-4 score was comparable to that of the Framingham risk score, whereas the Bard score exhibited the worst performance. In the KoGES cohort, the GPT-4 score had the highest accuracy of 0·902 and specificity of 0·926. Table 3 details the clinical examples of the participants’ data, Framingham risk scores calculated using the data, and risk scores derived from LLMs. Table 4 shows the LLM model performance using limited information. GPT-3·5’s prediction performance was preserved despite omitting a few variables in the prompt, particularly without physical measurement data.

**Table 2.** Performance comparison of Framingham, Bard, and ChatGPT Risk Score

|  | Accuracy | Sensitivity | Specificity | PPV | NPV | F1 score |
| --- | --- | --- | --- | --- | --- | --- |
| **UK biobank** |  |  |  |  |  |  |
| GPT-4 | **0.834** | 0.393 | **0.849** | **0.084** | 0.975 | **0.138** |
| GPT-3.5 | 0.674 | **0.598** | 0.677 | 0.061 | **0.980** | 0.111 |
| Bard | 0.702 | 0.447 | 0.711 | 0.052 | 0.973 | 0.093 |
| Framingham | 0.773 | 0.508 | 0.782 | 0.076 | 0.978 | 0.132 |
| **KoGES** |  |  |  |  |  |  |
| GPT-4 | **0.902** | 0.153 | **0.926** | 0.062 | 0.972 | 0.088 |
| GPT-3.5 | 0.836 | 0.273 | 0.854 | 0.056 | 0.974 | 0.093 |
| Bard | 0.779 | 0.307 | 0.794 | 0.045 | 0.973 | 0.079 |
| Framingham | **0.874** | **0.278** | 0.893 | **0.077** | **0.975** | **0.120** |

PPV: positive predictive value, NPV: negative predictive value. Bold font indicates the highest value of the corresponding metric.

**Table 3.**
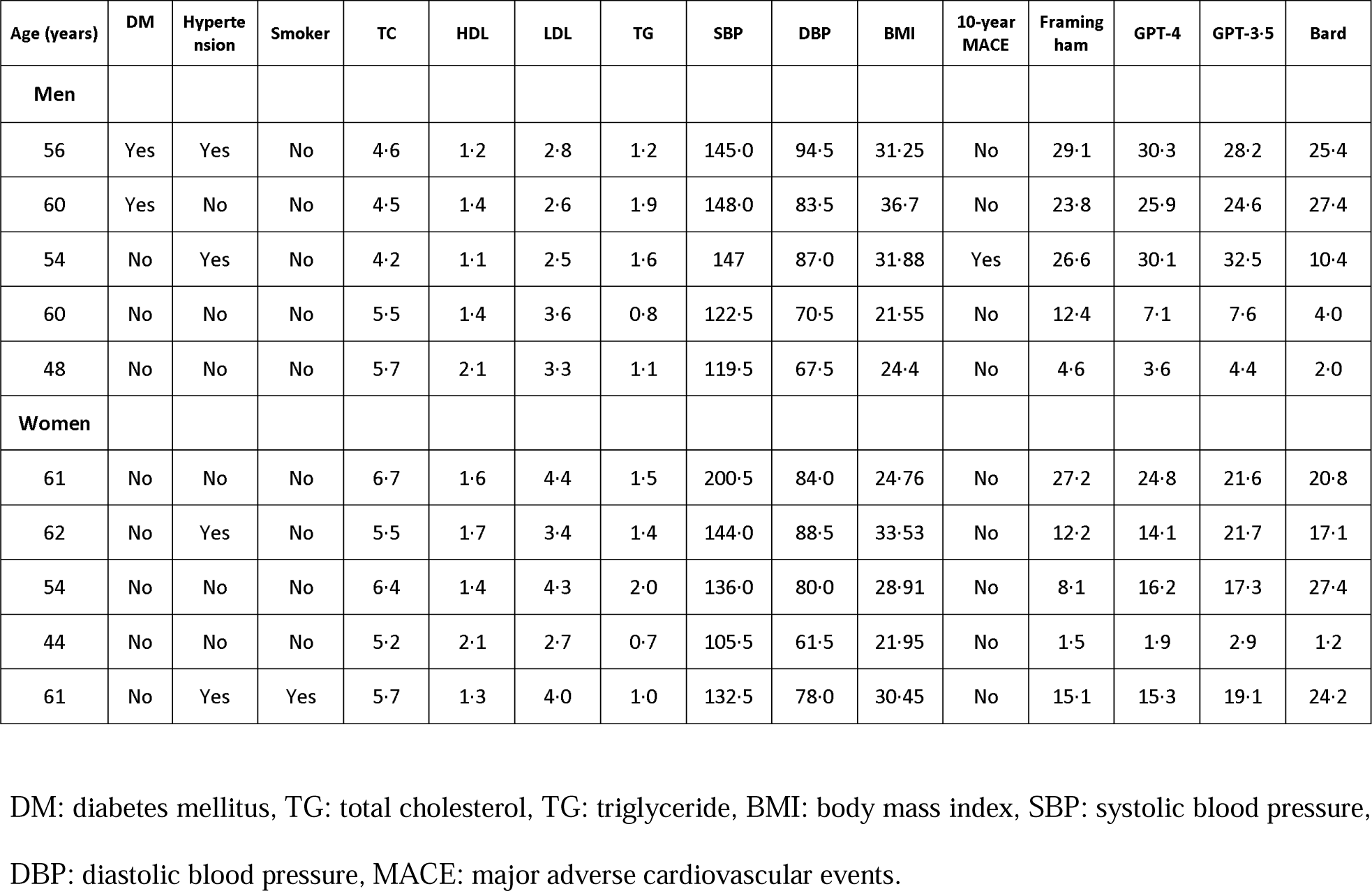
Examples of participants’ data and the scores derived from LLMs

**Table 4.** LLM model performance using limited information

|  | Accuracy | Sensitivity | Specificity | PPV | NPV | F1 score |
| --- | --- | --- | --- | --- | --- | --- |
| **GPT-3.5 baseline** | 0.674 | 0.598 | 0.677 | 0.061 | 0.98 | 0.111 |
| **Omitting** |  |  |  |  |  |  |
| **Medical history** | 0.5 | 0.719 | 0.493 | 0.048 | 0.98 | 0.089 |
| **Lipid profile** | 0.653 | 0.583 | 0.656 | 0.056 | 0.978 | 0.103 |
| **Physical measurement** | 0.712 | 0.505 | 0.719 | 0.059 | 0.976 | 0.106 |

“Medical history” in the omitting section is defined as a prompt that excludes a patient’s medical history information, such as smoking status, diabetes, and hypertension, from the existing prompt. “Lipid profile” is defined as a prompt that excludes the patient’s lipid profile from the existing prompt. “Physical measurement” is defined as a prompt that excludes a patient's physical measurements, such as blood pressure and BMI, from the existing prompt. PPV: positive predictive value, NPV: negative predictive value.

Fig. 3 displays scatterplots and Pearson correlation coefficients (Pearson’s r) for the different scoring systems. GPT-4 had the highest correlation with the Framingham risk score (Pearson’s r = 0·753), followed by GPT-3·5 (Pearson’s r = 0·709), and Bard (Pearson’s r = 0·446). Pearson’s r between GPT-4 and GPT-3·5 was 0·626.

**Fig. 3.**
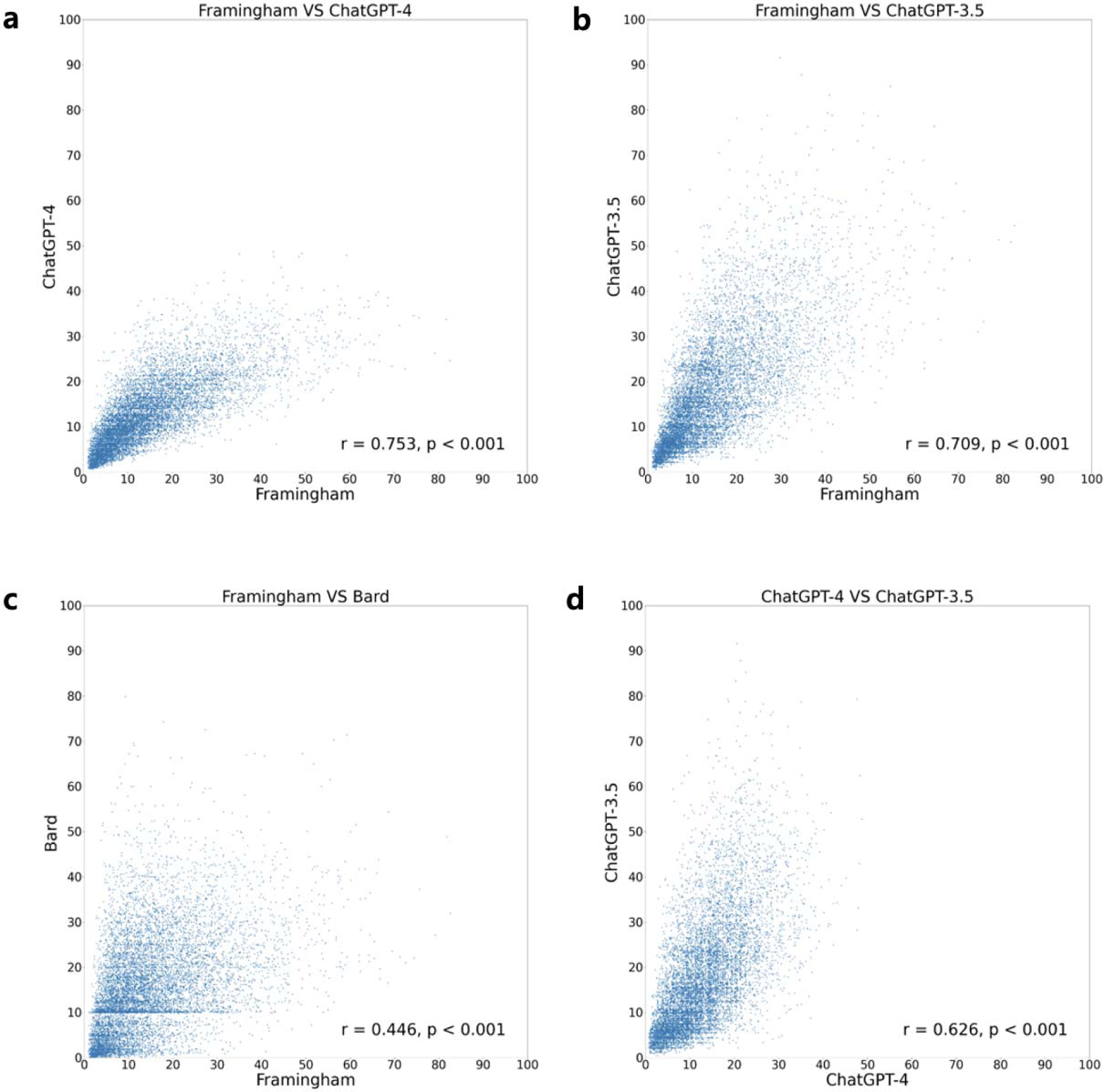
Scatterplots and Pearson correlation coefficient for various scoring methods. GPT-4 and Framingham risk score exhibit satisfactory correlation between each other. All pairs show statistically significant correlation.

Fig. 4 displays the Kaplan–Meier curves stratified by risk using the different scoring systems. All pairwise comparisons (with or without Bonferroni correction) between curves using the log-rank test were statistically significant. Fig. 5 and Supplementary Table 3 show the hazard ratios (HRs) for 10-year MACE of the moderate- and high-risk groups compared to the low-risk group in each scoring system using the Cox proportional hazards model. The HRs of GPT-4 were comparable to that of the Framingham risk score (GPT-4 moderate risk HR 2·94, 95% CI 2·15-4·02, GPT-4 high risk HR 6·81, 95% CI 4·96-9·36, Framingham moderate risk HR 3·17, 95% CI 2·27-4·45, Framingham high risk HR 6·96, 95% CI 5·05-9·60). The HRs of GPT-3·5 were 2·44 (95% CI 1·70-3·50) for moderate risk and 5·05 (95% CI 3·64-7·00) for high risk, and the HRs of Bard were 1·80 (95% CI 1·32-2·47) for moderate risk and 2·84 (95% CI 2·09-3·87) for high risk.

**Fig. 4.**
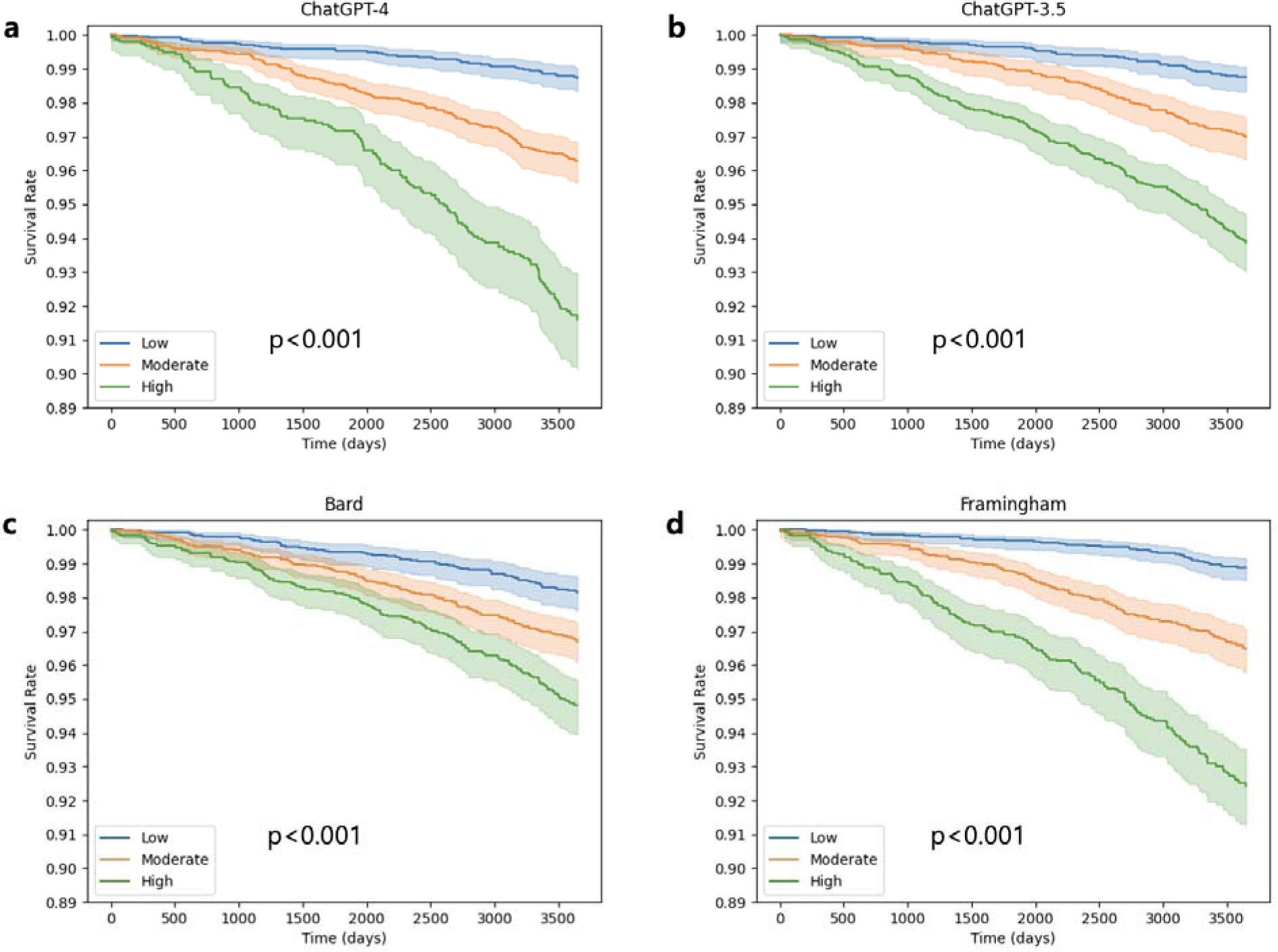
Kaplan–Meier curves stratified by cardiovascular risk on LLMs and Framingham risk scoring models. GPT-4 demonstrated distinct survival patterns among the groups, which revealed a strong association between the GPT’s risk prediction output and survival outcomes. All pairwise comparisons between curves in other models with the log-rank test were statistically significant.

**Fig. 5.**
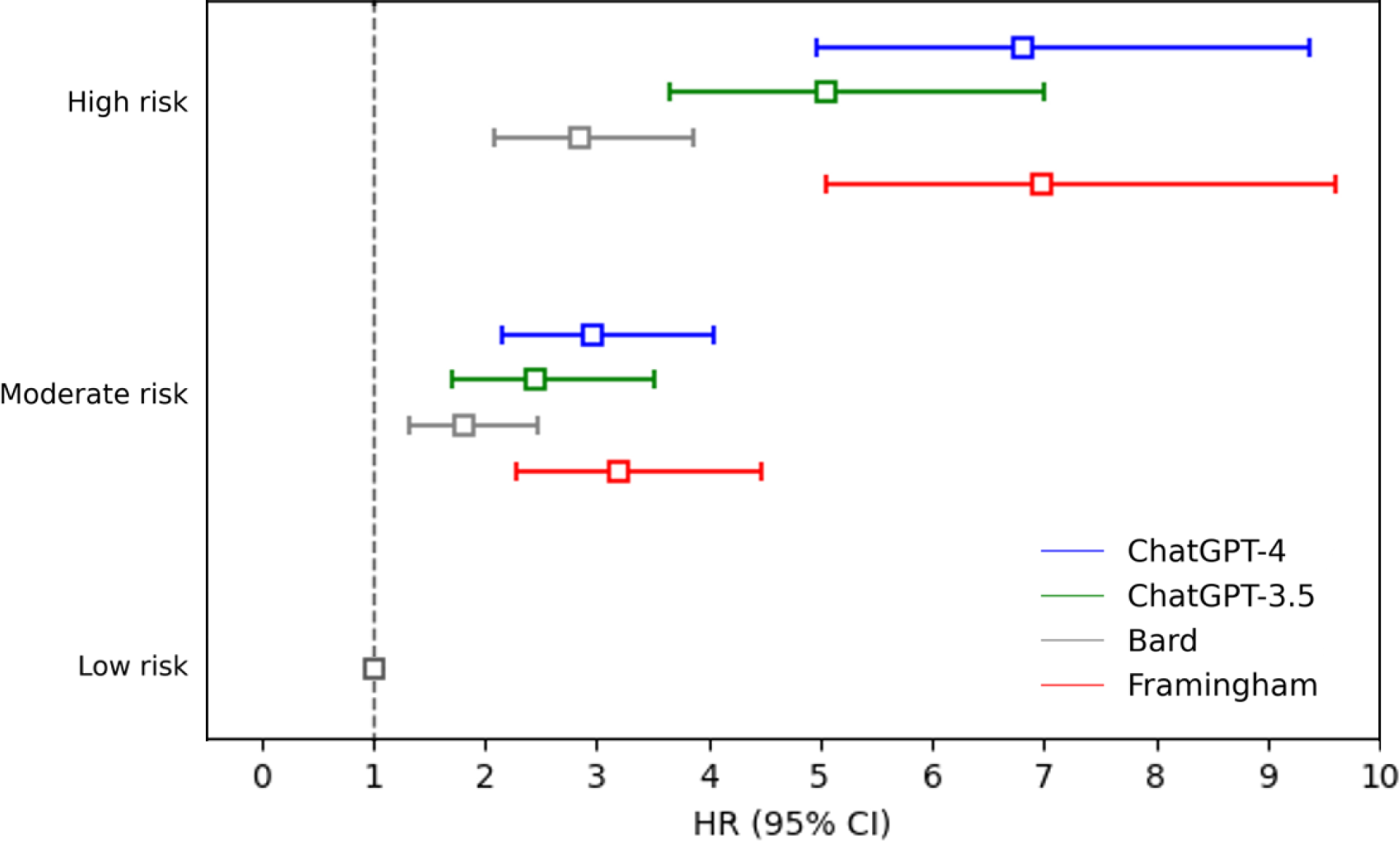
Comparison of hazard ratio between different scoring systems using Cox proportional hazards model. The hazard ratios for 10-year MACE of moderate risk and high risk were compared with the low-risk group in each scoring system using the cox proportional hazards model. The HRs of GPT-4 were comparable to that of the Framingham risk score. MACE, major adverse cardiovascular event; HR, hazard ratio; CI, confidence interval.

## Discussion

### Summary of findings

This study compared the performance of LLMs in cardiovascular risk prediction with that of the Framingham risk model and validated the output using real-world data. The findings of this study are as follows. GPT-4 achieved performance comparable to the FRS in cardiovascular risk prediction in both the UK Biobank {accuracy (0·834 vs· 0·773) and F1 score (0·138 vs· 0·132)} and KoGES {accuracy (0·902 vs· 0·874)}. The Kaplan–Meier survival analysis of GPT-4 demonstrated distinct survival patterns among groups, which revealed a strong association between the GPT risk prediction output and survival outcomes. The additional analysis of limited variables using GPT-3·5 indicated that ChatGPT’s prediction performance was preserved despite the omission of a few variables in the prompt, especially without physical measurement data (Table 4).

### Advantages for using LLM-based cardiovascular risk prediction

Since its release, ChatGPT has attracted considerable attention worldwide because of its exceptional ability to generate plausible responses across various topics. In some cases, ChatGPT has outperformed existing prediction models, encouraging studies on the potential of ChatGPT for use in various applications^11^. For instance, in the financial sector, compared with conventional analysis methods, ChatGPT has demonstrated superior performance in predicting stock prices^19^. However, limited studies have been conducted on the use of language models in healthcare. To the best of our knowledge, this study is the first to reveal that ChatGPT exhibited performance comparable with the conventional risk score model in predicting cardiovascular risk using large real-world medical data. These findings provide insights into the potential applicability of ChatGPT in medical practice.

Despite being a language-generation model, ChatGPT exhibits performance similar to the conventional model in predicting the cardiovascular risk of patients. Conventional prediction algorithms typically rely on multivariate regression of well-established CVD risk factors^3, 4^, typically limiting the number of risk factors and assuming linear relationships between them with minimal or no interaction between various factors^20^. By contrast, ChatGPT, which derives its answers by learning from large amounts of textual datasets to generate the most probable human-like responses rather than through mathematical calculations, achieves similar values and performances as linear computation-based regression models. Furthermore, conventional models frequently rely on old cohorts and have only been validated for specific cohort groups within individual countries, which limit their applicability to a broad population^21^. Furthermore, widely used calculators for renowned models such as the FRS have slightly distinct formulas based on different references, which results in heterogeneous prediction methods^22–24^. By contrast, ChatGPT can learn from multiple guidelines and select the most suitable guideline for prediction, which enhances its generalizability across a diverse population. ChatGPT has achieved satisfactory performance in the UK Biobank, Western database, KoGES, and Asian database, which highlights the generalizability and robustness of the model in diverse populations. ChatGPT provides improved accessibility and flexibility compared with conventional prediction models. Users can easily access ChatGPT by visiting a website, without requiring any particular application. The model rapidly delivered satisfactory outcomes without strict input constraints by accepting numerous input formats. ChatGPT can understand the context and range of the input values, even when precise units are not provided^25^. Unlike conventional prediction models, ChatGPT provides answers with limited input data. Because ChatGPT is a model that learns existing texts and derives results from them, cardiovascular risk can be determined from existing text data that may not contain the patient’s blood test results, medical history, or physical measurements. Therefore, unlike existing models, GPT can train any combination of variables and produce outputs through various types of input combinations. Furthermore, by estimating risks without requiring physical measurements such as height, weight, or systolic blood pressure data, ChatGPT predictions could potentially decrease hospital visits, enhance convenience, and promote advancements in telemedicine.

Conventional risk-stratification guidelines tend to be complex and require precise numerical values for each risk stratification, rendering the guidelines unsuitable for use in brief outpatient settings. However, with ChatGPT, users can request outputs in a specific format, which allows faster interpretation of medical records. For instance, users can rapidly access only the necessary information by asking ChatGPT to provide a patient’s cardiovascular risk score, utilized guidelines, and corresponding risk group.

### Future impact on LLM-based research

The potential benefits of ChatGPT, as demonstrated in the present analysis, may facilitate large-scale and adaptable cardiovascular risk assessments in the general population in the near future. The findings of this study indicate that ChatGPT can compute cardiovascular risk with reasonable accuracy using only facts expressed in natural language, even in the absence of certain data. Consequently, this approach facilitates the monitoring of CVD risk in a larger population, which promotes earlier interventions and management of at-risk patients. Moreover, the features of ChatGPT observed in this study have considerable implications in both clinical practice and research. ChatGPT can enable the semantic extraction of targets of interest. The operational definitions for determining the study subjects vary across institutions and studies^26^. Therefore, selecting patients with consistent meanings from various institutions using conventional methods is challenging given the discrepancies in data formats and meanings. However, ChatGPT can function independently of data formats and adherence to standards. Thus, by converting existing data into text, semantically extracting the patients of interest becomes feasible. This approach facilitated the integration and analysis of disparate data from multiple institutions. For example, by leveraging the findings of this study, individuals with specific levels of cardiovascular risk can be promptly identified.

Finally, this study has a few limitations. First, the unavailability of API for GPT-4 and Bard limited our analysis to a subset of 10,000 UK Biobank participants. However, this result was sufficient to validate the findings of this study. Second, the inner workings of GPT-4 remain challenging because the model and the code of ChatGPT have not been fully disclosed, and because of the complex structure of LLM, fully explaining the working principle becomes difficult. Third, the performance of GPT-4 is yet to be extensively validated for various medical conditions, necessitating additional research to generalize our findings to other conditions such as diabetes or cancer. Studies are required to optimize the performance of GPT-4 through fine-tuning and prompt engineering of specific tasks.

LLMs, such as ChatGPT, can achieve performance comparable to conventional models in predicting cardiovascular risk. Furthermore, ChatGPT exhibits enhanced accessibility, flexibility, and ability to provide user-friendly outputs. With the continuous evolution of LLMs, such as ChatGPT, future studies should focus on applying the models to various medical scenarios and optimizing their performance.

## Data Availability

All data produced in the present work are contained in the manuscript.

## Contributors

Conceptualization: SungA Bae,

Methodology: Dukyong Yoon,

Validation: Seng Chan You,

Investigation: Changho Han, Dong Won Kim, Songsoo Kim,

Writing—original draft preparation: Changho Han, Dong Won Kim, Songsoo Kim,

Writing—review and editing: Dukyong Yoon, SungA Bae, Seng Chan You,

Supervision: Dukyong Yoon, SungA Bae

## Data sharing

Deidentified data, from both the UK biobank and KoGES studies, is publicly available on request. Further details of UK biobank and KoGES studies are available online.

## Acknowledgements

This study was conducted using data from the UK Biobank (application number: 85037). Data in this study were from the Korean Genome and Epidemiology Study (KOGES; 6635-302), National Research Institute of Health, Korea Disease Control and Prevention Agency, Republic of Korea.

