## Supplementary Materials for "Large-language-model-based 10-year risk prediction of cardiovascular disease: insight from the UK biobank data"

**Supplementary Table 1 | Summary of UK biobank data**

This table encompasses the official names (descriptions) and unique field numbers for each variable from the UK Biobank and denotes the value type of each variable as an integer, categorical, continuous, or date. The table outlines the number of participants surveyed for each variable.

| **UK biobank data field** | | | |
| --- | --- | --- | --- |
| **Description** | **Field No.** | **Value Type** | **Participants** |
| Sex | 31 | Categorical | 502,371 |
| Standing height | 50 | Continuous | 499,830 |
| Date of attending assessment centre | 53 | Date | 502,371 |
| Diabetes diagnosed by doctor | 2443 | Categorical | 501,438 |
| Diastolic blood pressure | 4079 | Integer | 472,259 |
| Systolic blood pressure | 4080 | Integer | 472,254 |
| Medication for cholesterol, blood pressure, diabetes, or take exogenous hormones | 6153 | Categorical | 270,772 (Female only) |
| Medication for cholesterol, blood pressure, or diabetes | 6177 | Categorical | 226,881 (Male only) |
| Smoking status | 20116 | Categorical | 501,478 |
| Ethnic background | 21000 | Categorical | 501,479 |
| Weight | 21002 | Continuous | 499,594 |
| Age at recruitment | 21022 | Integer | 502,368 |
| Cholesterol | 30680 | Continuous | 469,467 |
| HDL cholesterol | 30760 | Continuous | 429,761 |
| LDL direct | 30780 | Continuous | 468,584 |
| Triglycerides | 30870 | Continuous | 469,092 |
| Date of death | 40000 | Date | 44,559 |

**Supplementary Table 2 | Summary of KoGES data**

The table lists variables under categories and the values corresponding to each baseline variable name. The value types include date, integer, categorical, and float, with the number of participants for each variable provided in the respective column. For LDL calculations, total cholesterol, HDL, and triglycerides were used as the basis and incorporated into the analysis.

| **KoGES data** | | | | |
| --- | --- | --- | --- | --- |
| **Table name** | **Description** | **Baseline Variable Name** | **Value Type** | **Participants** |
| MM2_01_GEN | investigation date | A01_EDATE | Date | 10,030 |
| MM2_01_GEN | Sex | A00_SEX | Categorical | 10,030 |
| MM2_01_GEN | Age | A01_AGE | Integer | 10,030 |
| MM2_03_SMOKE | Smoking status | A01_SMOKE | Categorical | 9,896 |
| MM2_05_DRUG_1 | Medication for blood pressure | A01_DRUGHT | Categorical | 9,445 |
| MM2_06_DISEASE_C | Diagnosis of diabetes | A01_DM_C | Categorical | 10,026 |
| MM2_07_DISEASE_1 | Diagnosis of Myocardial infarction | A01_MI | Categorical | 10,026 |
| MM2_07_DISEASE_2 | Diagnosis of cerebrovascular disease (stroke, etc.) | A01_CEVA | Categorical | 10,026 |
| MM2_14_BIOCHEM_2 | Total cholesterol | A01_TCHL_ORI | float | 10,028 |
| MM2_14_BIOCHEM_2 | HDL | A01_HDL_ORI | float | 10,028 |
| MM2_14_BIOCHEM_2 | Triglycerides | A01_TRIGLY_ORI | float | 10,027 |
| MM2_15_ANTHRO | Height | A01_HEIGHT | float | 10,030 |
| MM2_15_ANTHRO | Weight | A01_WEIGHT | float | 10,026 |
| MM2_15_ANTHRO | Left arm SBP | A01_SBP_L | float | 10,030 |
| MM2_15_ANTHRO | Right arm SBP | A01_SBP_R | float | 10,030 |
| MM2_15_ANTHRO | Left arm DBP | A01_DBP_L | float | 10,030 |
| MM2_15_ANTHRO | Right arm DBP | A01_DBP_R | float | 10,030 |

**Supplementary Table 3 | Hazard ratios for 10-year MACE of the moderate- and high-risk groups compared to the low-risk group in each scoring system using the Cox proportional hazards model.**

|  | **GPT-4** | | |  | **GPT-3·5** | | |  | **Bard** | | |  | **Framingham** | | |
| --- | --- | --- | --- | --- | --- | --- | --- | --- | --- | --- | --- | --- | --- | --- | --- |
|  | **HR** | **95% CI** | **p-value** |  | **HR** | **95% CI** | **p-value** |  | **HR** | **95% CI** | **p-value** |  | **HR** | **95% CI** | **p-value** |
| **Moderate risk** | 2·94 | 2·15-4·02 | <0·001 |  | 2·44 | 1·70-3·50 | <0·001 |  | 1·80 | 1·32-2·47 | <0·001 |  | 3·17 | 2·27-4·45 | <0·001 |
| **High risk** | 6·81 | 4·96-9·36 | <0·001 |  | 5·05 | 3·64-7·00 | <0·001 |  | 2·84 | 2·09-3·87 | <0·001 |  | 6·96 | 5·05-9·60 | <0·001 |

**Supplementary figure 1 | Flowchart of KoGES study population selection**

The KoGES dataset comprises 10,030 participants, who were enrolled based on baseline surveys conducted between 2001 and 2002 in the cities of Ansan and Anyang. In our analysis, we excluded 875 individuals with at least one missing value in the variables such as smoking status, medication for blood pressure, diagnosis of myocardial infarction, diagnosis of cerebrovascular disease, total cholesterol, HDL, triglycerides, and weight. Consequently, we focused on a cohort of 9,155 individuals. Additionally, the dataset was utilized to conduct follow-up assessments over 10 years to monitor the occurrence of myocardial infarction or stroke. After excluding 3,437 individuals who were not followed up at the 10-year mark (5th follow-up), the study proceeded with a final sample of 5,718 participants.

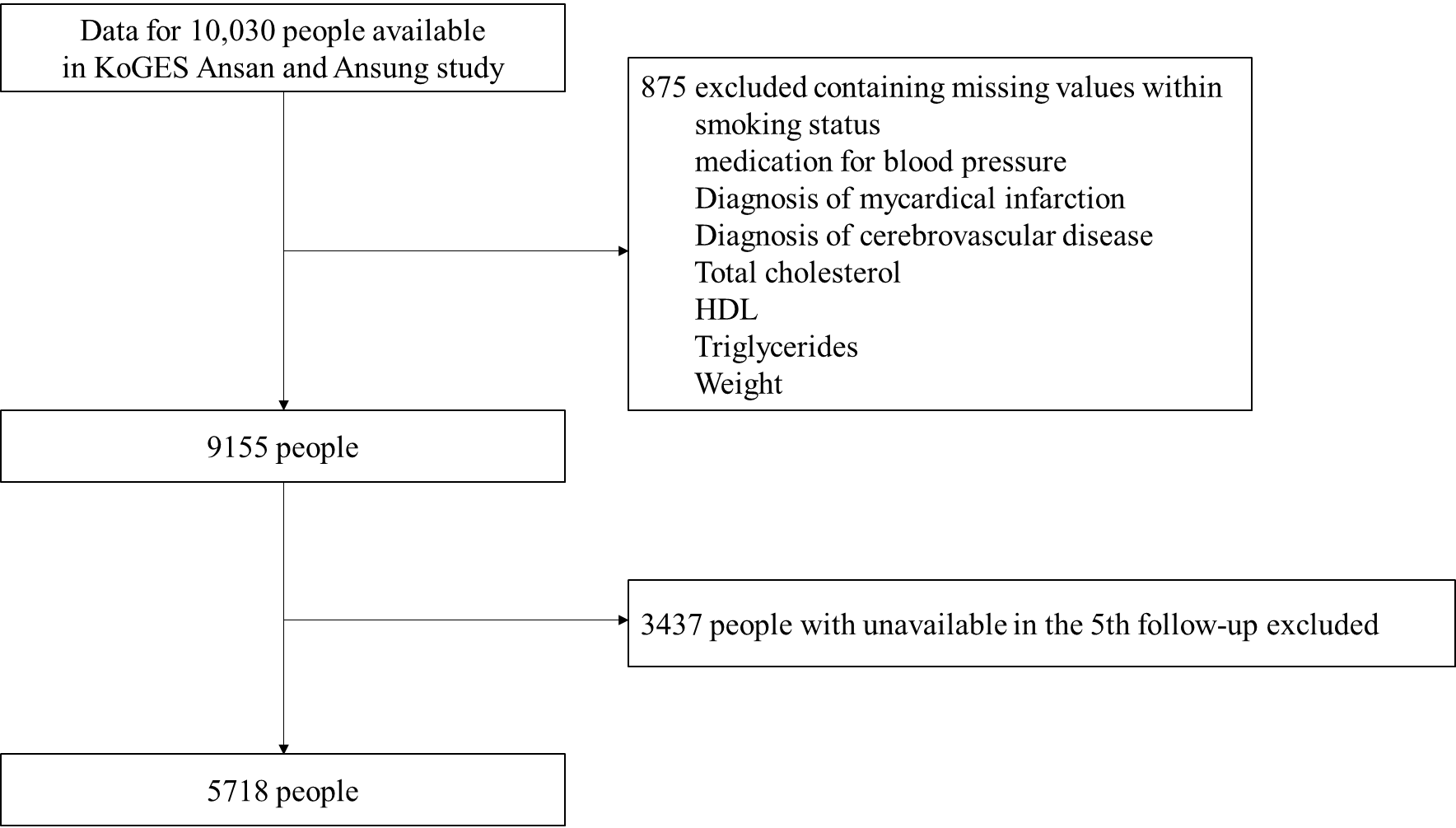
